# Expanding the Pediatric Heart Donor Pool: National Outcomes of Donation After Circulatory Death Versus Donation After Brain Death Heart Transplantation

**DOI:** 10.64898/2026.07.03.26357254

**Authors:** Bilal Khan Mohammed, Rohit Ganduboina, Omar Abdel Kerim, Gayatri Muley, Palak Dutta, Nitya Krishna Arumugam, John Karamichalis, Yahiya Pasha Syed, Sandeep Sainathan

**Author notes:** **Corresponding author:** Bilal Khan Mohammed, MD, Division of Cardiac Surgery, Northwestern University Feinberg School of Medicine, Chicago, IL, USA. Telephone: []. Bilal Khan Mohammed and Rohit Ganduboina contributed equally to this work and are designated co-first authors. **Funding:** None. **Data availability:** Data were obtained from the OPTN/UNOS Standard Transplant Analysis and Research files and are available to qualified investigators through the OPTN. **Ethics:** This study used de-identified registry data and was deemed exempt from institutional review board review. **Prior presentation:** Presented at the Eastern Cardiothoracic Surgical Society (ECTSS) Annual Meeting, 2025 (Abstract C02).

## Abstract

**Background:** Donation after circulatory death (DCD) is an increasingly accepted strategy to expand the adult heart donor pool, but its use in children remains limited and incompletely characterized. We compared national characteristics and post-transplant outcomes of pediatric DCD versus donation after brain death (DBD) heart transplantation.

**Methods:** We performed a retrospective cohort study of the Organ Procurement and Transplantation Network (OPTN) registry, including patients younger than 18 years who underwent primary isolated heart transplantation between January 1993 and March 2025. Recipients were stratified by donor type (DCD vs DBD). Continuous variables were compared with the Mann–Whitney U test and categorical variables with the χ^2^ or Fisher exact test. Survival was estimated by the Kaplan–Meier method and compared using the log-rank test and Cox proportional hazards regression.

**Results:** Of 10,671 pediatric heart transplant recipients, 33 (approximately 0.3%) received DCD allografts. The first DCD transplant was recorded in 2004, with a marked increase in 2023–2024. Compared with DBD recipients, DCD recipients were more frequently infants (<1 year, 51.5% vs 28.4%) and more often had congenital heart disease (69.7% vs 47.6%; P=0.033); DCD donors were younger (median 0 vs 6 years; P=0.038) and more frequently died of anoxia (72.7% vs 37.0%; P<0.001). Donor and recipient left ventricular mass were lower in the DCD group (P<0.05), but predicted left ventricular mass matching was similar. DCD recipients had longer hospital stays (median 31.5 vs 19 days; P=0.023); rates of treated rejection, dialysis, stroke, and pacemaker implantation were comparable. Early survival did not differ (30-day, 90-day, and 1-year), and Kaplan–Meier survival through 5 years was not significantly different (hazard ratio 1.17; 95% CI 0.49–2.81; log-rank P=0.73). More than 90% of DCD transplants were performed in four UNOS regions (11, 4, 5, and 8).

**Conclusions:** In this national analysis, pediatric DCD heart transplantation was uncommon but expanding rapidly, concentrated in a few regions, and used preferentially in infants and children with congenital heart disease. Early post-transplant outcomes were not significantly different from DBD, supporting cautious expansion of DCD as a means of enlarging the pediatric donor pool. The small number of DCD recipients and limited follow-up warrant confirmation in larger, longer-term studies.

## Introduction

Heart transplantation remains the definitive therapy for children with end-stage heart failure, yet its reach is constrained by a persistent shortage of suitable donor organs. Waitlist mortality remains substantial, particularly among infants and children with congenital heart disease, in whom size-matched donor availability is most limited [1]. Strategies that safely enlarge the donor pool therefore have the potential to reduce waitlist death and expand access to transplantation.

Donation after circulatory death (DCD) has re-emerged as one such strategy. Although the first pediatric DCD heart transplants were reported in 2008 using direct procurement [2], contemporary adult experience was catalyzed by the 2015 report of distant DCD procurement with ex-situ machine perfusion [3] and by subsequent demonstrations that hearts recovered after circulatory death could be functionally assessed and successfully transplanted [4]. The first randomized trial comparing DCD with DBD heart transplantation demonstrated non-inferior early survival [5], accelerating adoption. National analyses in adults have shown that DCD shortens waiting times without compromising post-transplant outcomes [6,7].

Experience in children has lagged. The largest early pediatric series, drawn from the International Society for Heart and Lung Transplantation registry, identified only 21 DCD transplants and suggested lower early survival, likely reflecting an early learning curve [8]. More recently, the first United States pediatric DCD heart transplant using ex-vivo perfusion was reported [9], and normothermic regional perfusion has been introduced as an alternative recovery strategy suited to smaller donors [10,11]. National registry analyses have begun to describe increasing pediatric DCD utilization [12] and improved waitlist outcomes among candidates willing to accept DCD organs [13], although data in adults with congenital heart disease have raised the possibility of worse graft survival in selected populations [14]. Despite this growing interest, contemporary national data comparing post-transplant outcomes between DCD and DBD recipients across the pediatric age spectrum remain limited.

We therefore used the OPTN registry to characterize the utilization, recipient and donor characteristics, geographic distribution, and post-transplant outcomes of pediatric DCD heart transplantation compared with DBD, with the goal of informing the safe expansion of the pediatric donor pool.

## Methods

### Data source and study population

We conducted a retrospective cohort study using the Organ Procurement and Transplantation Network (OPTN) registry, which prospectively captures recipient, donor, waitlist, and transplant-related data from all United States transplant centers and has been extensively used for pediatric heart transplantation research [15]. We included patients younger than 18 years who underwent primary isolated heart transplantation between January 1993 and March 2025. Because pediatric DCD heart transplantation was first recorded in the OPTN registry in 2004, all eligible DCD recipients from 2004 onward were included, whereas DBD recipients were identified throughout the study period. Patients undergoing repeat transplantation, multiorgan transplantation, or with missing donor-type information were excluded. The final cohort comprised 10,671 recipients (33 DCD and 10,638 DBD).

### Variables and outcomes

Recipient variables included age, sex, race, body mass index, blood type, heart failure etiology, prior cardiac surgery, cerebrovascular disease, renal and hepatic function, cytomegalovirus serostatus, pre-transplant transfusion, mechanical ventilation, inotropic support, and invasive hemodynamics (cardiac index, mean pulmonary artery pressure, pulmonary capillary wedge pressure, pulmonary vascular resistance, and transpulmonary gradient). Donor variables included age, sex, race, body mass index, blood type, mechanism of death, serum creatinine, total bilirubin, donor–recipient weight and height mismatch, HLA mismatch, donor distance to the transplant center, ischemic time, and donor and recipient predicted left ventricular mass (PLM) with the PLM ratio. Transplant variables included transplant year, waitlist duration, listing status, and pre-transplant mechanical circulatory support.

The primary outcome was post-transplant all-cause mortality. Secondary outcomes included 30-day, 90-day, 1-year, and 5-year survival, dialysis, treated acute rejection, stroke, permanent pacemaker implantation, hospital length of stay, temporal trends in DCD utilization, and geographic variation across the 11 United Network for Organ Sharing (UNOS) regions.

### Statistical analysis

Analyses were performed using complete-case data without imputation, in Python (version 3.13). Continuous variables are reported as medians with interquartile ranges (IQR) and compared with the Mann–Whitney U test; categorical variables are reported as frequencies with percentages and compared with the Pearson χ ^2^ or Fisher exact test, as appropriate. Post-transplant survival was estimated by the Kaplan–Meier method and compared with the log-rank test, and Cox proportional hazards regression was used to estimate the hazard ratio (HR) with 95% confidence interval (CI) for donor type. Given the small number of DCD events, the donor-type comparison was not adjusted in a multivariable model, and the corresponding estimate should be regarded as unadjusted. Temporal trends were evaluated by transplant year, and geographic variation was assessed as the proportion of pediatric DCD transplants within each UNOS region. A two-sided P value <0.05 was considered statistically significant.

## Results

### Utilization and temporal trends

Among 10,671 pediatric heart transplant recipients, 33 (approximately 0.3%) received allografts from DCD donors. The first pediatric DCD heart transplant was recorded in 2004, after which annual use remained negligible until a pronounced increase in 2023–2024 (Figure 1). Consistent with this, the median transplant year was markedly later for DCD than DBD recipients (2023 [IQR 2023–2024] vs 2013 [IQR 2004–2019]; P<0.001).

**Figure 1.**
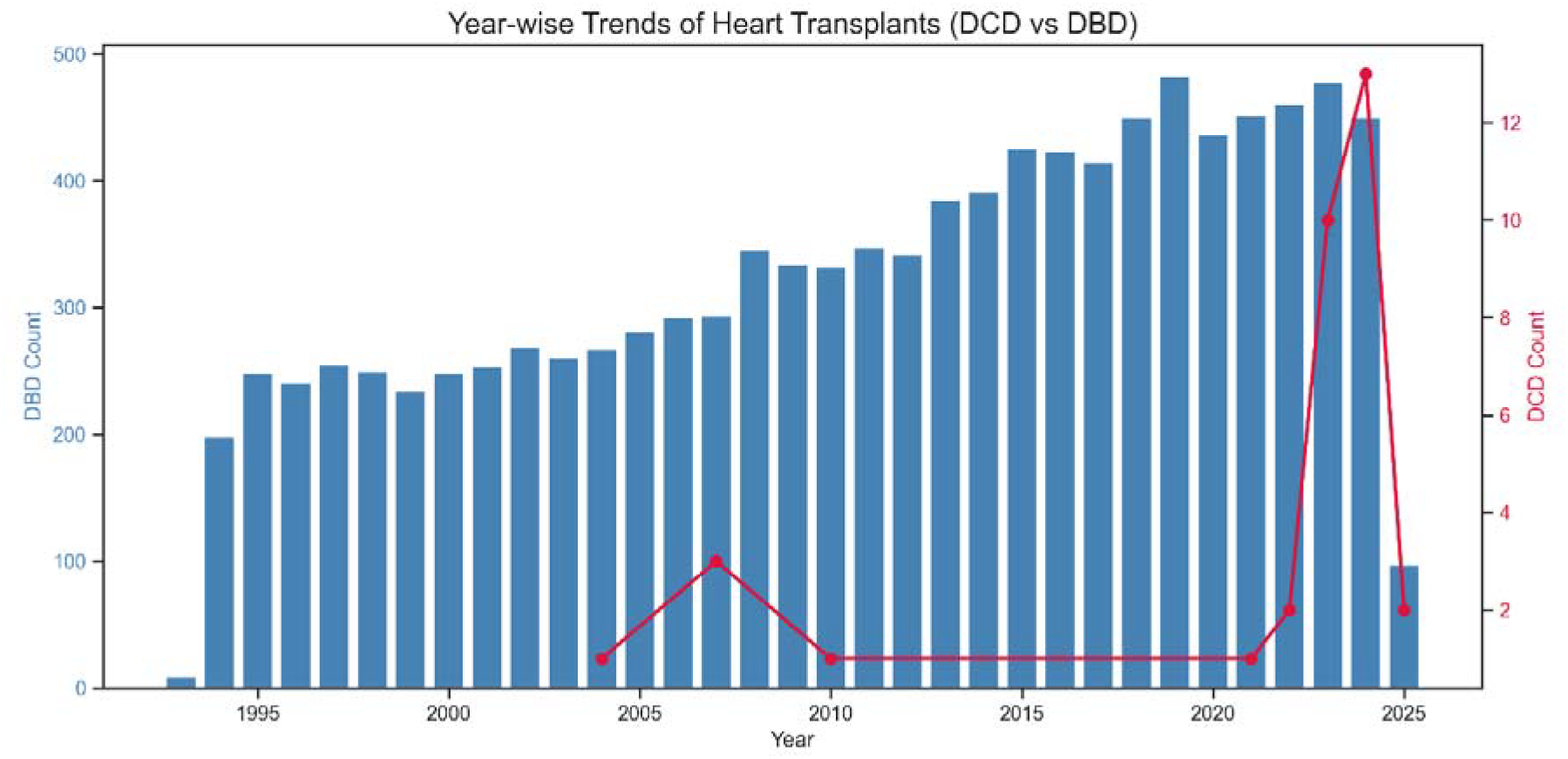
Annual utilization of pediatric heart transplantation by donor type, 1993–2025. Bars indicate the number of donation after brain death (DBD) transplants (left axis); the line indicates the number of donation after circulatory death (DCD) transplants (right axis). DCD transplants first appear in 2004 and increase markedly in 2023–2024.

### Recipient and donor characteristics

DCD recipients were more frequently infants than DBD recipients (<1 year, 51.5% vs 28.4%; age-group P=0.012) and more often had congenital heart disease (69.7% vs 47.6%; P=0.033), with correspondingly less cardiomyopathy. Sex, race, body mass index, blood type, and pre-transplant hemodynamic and support characteristics did not differ significantly between groups (Table 1). DCD donors were younger (median 0 [IQR 0–13] vs 6 [IQR 1–16] years; P=0.038) and more frequently died of anoxia (72.7% vs 37.0%), whereas head trauma predominated among DBD donors (mechanism of death P<0.001).

**Table 1.** Recipient, donor, and transplant characteristics and post-transplant outcomes of pediatric DCD versus DBD heart transplant recipients.

| Variable | DBD (n=10,638) | DCD (n=33) | P value |
| --- | --- | --- | --- |
| **Transplant characteristics** |  |  |  |
| Total | 10,638 | 33 |  |
| **Recipient characteristics** |  |  |  |
| Age group, n (%) |  |  | **0.012** |
| <1 year | 3024 (28.4) | 17 (51.5) |  |
| 1–5 years | 2497 (23.5) | 4 (12.1) |  |
| 6–10 years | 1452 (13.6) | 6 (18.2) |  |
| 11–17 years | 3665 (34.5) | 6 (18.2) |  |
| Female, n (%) | 4655 (43.8) | 14 (42.4) | 1.000 |
| Race, n (%) |  |  | 0.567 |
| Caucasian | 5869 (55.2) | 13 (39.4) |  |
| Hispanic | 1982 (18.6) | 8 (24.2) |  |
| African American | 2089 (19.6) | 8 (24.2) |  |
| Asian | 397 (3.7) | 2 (6.1) |  |
| Other | 282 (2.7) | 1 (3.0) |  |
| Body mass index (kg/m<sup>2</sup>) | 16.76 [14.79–19.71] | 15.71 [13.89–18.37] | 0.085 |
| Blood type, n (%) |  |  | 0.068 |
| O | 4833 (45.4) | 22 (66.7) |  |
| A | 3942 (37.1) | 6 (18.2) |  |
| B | 1414 (13.3) | 3 (9.1) |  |
| AB | 449 (4.2) | 2 (6.1) |  |
| Heart failure etiology, n (%) |  |  | **0.033** |
| Congenital heart disease | 5067 (47.6) | 23 (69.7) |  |
| Cardiomyopathy | 5191 (48.8) | 10 (30.3) |  |
| Other | 380 (3.6) | 0 (0.0) |  |
| Prior cerebrovascular accident, n (%) | 303 (2.8) | 2 (6.1) | 0.580 |
| Prior cardiac surgery, n (%) | 526 (4.9) | 2 (6.1) | 1.000 |
| Prior dialysis, n (%) | 268 (2.5) | 1 (3.0) | 1.000 |
| Total bilirubin (mg/dL) | 0.60 [0.40–1.20] | 0.60 [0.40–0.80] | 0.859 |
| Serum creatinine (mg/dL) | 0.47 [0.30–0.70] | 0.50 [0.30–0.68] | 0.616 |
| Positive CMV serology, n (%) | 3378 (31.8) | 10 (30.3) | 0.714 |
| Blood transfusion on waitlist, n (%) | 4317 (40.6) | 14 (42.4) | 1.000 |
| Pre-transplant mechanical ventilation, n (%) | 3331 (31.3) | 17 (51.5) | 0.098 |
| Intravenous inotropes, n (%) | 5049 (47.5) | 16 (48.5) | 1.000 |
| Cardiac index (L/min/m<sup>2</sup>) | 2.77 [2.12–3.68] | 2.30 [2.17–3.03] | 0.382 |
| Mean pulmonary artery pressure (mm Hg) | 22.0 [17.0–31.0] | 24.0 [20.0–30.3] | 0.431 |
| Pulmonary capillary wedge pressure (mm Hg) | 15.0 [10.0–21.0] | 14.0 [14.0–16.0] | 0.750 |
| Pulmonary vascular resistance (Wood units) | 2.58 [1.42–4.82] | 5.13 [1.23–6.63] | 0.602 |
| Transpulmonary gradient (mm Hg) | 7.0 [5.0–11.0] | 8.0 [5.0–10.0] | 0.898 |
| **Donor characteristics** |  |  |  |
| Age (years) | 6.0 [1.0–16.0] | 0.0 [0.0–13.0] | **0.038** |
| Female, n (%) | 4318 (40.6) | 16 (48.5) | 0.457 |
| Race, n (%) |  |  | 0.765 |
| Caucasian | 5916 (55.6) | 21 (63.6) |  |
| African American | 2287 (21.5) | 5 (15.2) |  |
| Hispanic | 2095 (19.7) | 6 (18.2) |  |
| Asian | 181 (1.7) | 1 (3.0) |  |
| Other | 151 (1.4) | 0 (0.0) |  |
| Body mass index (kg/m<sup>2</sup>) | 18.54 [16.01–22.12] | 18.53 [14.69–22.12] | 0.218 |
| Blood type, n (%) |  |  | 0.119 |
| O | 6233 (58.6) | 26 (78.8) |  |
| A | 3254 (30.6) | 6 (18.2) |  |
| B | 977 (9.2) | 1 (3.0) |  |
| AB | 174 (1.6) | 0 (0.0) |  |
| Mechanism of death, n (%) |  |  | <0.001 |
| Anoxia | 3937 (37.0) | 24 (72.7) |  |
| Head trauma | 5498 (51.7) | 9 (27.3) |  |
| Cerebrovascular accident | 758 (7.1) | 0 (0.0) |  |
| CNS tumor | 76 (0.7) | 0 (0.0) |  |
| **Size matching and transplant characteristics** |  |  |  |
| Weight mismatch >±20%, n (%) | 6575 (61.8) | 24 (72.7) | 0.267 |
| Height mismatch >±5%, n (%) | 7577 (71.2) | 22 (66.7) | 0.700 |
| HLA mismatch >3 loci, n (%) | 7789 (73.2) | 23 (69.7) | 0.795 |
| Donor distance to center (nautical miles) | 297 [126–453] | 465 [201–599] | 0.056 |
| Ischemic time (hours) | 3.58 [2.97–4.25] | 3.72 [2.30–4.79] | 0.558 |
| Status at transplantation, n (%) |  |  | 0.215 |
| Status 1A | 7654 (71.9) | 24 (72.7) |  |
| Status 1B | 1203 (11.3) | 7 (21.2) |  |
| Status 2 | 907 (8.5) | 2 (6.1) |  |
| Old Status 1 | 872 (8.2) | 0 (0.0) |  |
| Temporarily inactive | 2 (0.0) | 0 (0.0) |  |
| Donor LV mass (g) | 45.17 [21.87–107.58] | 21.34 [9.53–92.05] | **0.020** |
| Recipient LV mass (g) | 34.50 [15.97–78.67] | 19.07 [9.38–44.51] | **0.004** |
| Predicted LV mass ratio | 1.35 [1.08–1.93] | 1.36 [1.11–2.04] | 0.765 |
| PLM ratio category, n (%) |  |  | 0.941 |
| >0.90 | 9868 (92.8) | 30 (90.9) |  |
| ≤0.90 | 770 (7.2) | 3 (9.1) |  |
| Transplant year, median [IQR] | 2013 [2004–2019] | 2023 [2023–2024] | <0.001 |
| Days on waitlist | 53 [19–128] | 44 [6–111] | 0.363 |
| **Post-transplant outcomes** |  |  |  |
| 30-day survival, n (%) | 10,221 (96.1) | 33 (100.0) | 0.477 |
| 90-day survival, n (%) | 9986 (93.9) | 32 (97.0) | 0.706 |
| 1-year survival, n (%) | 9591 (90.2) | 29 (87.9) | 0.884 |
| Treated acute rejection, n (%) | 1958 (18.4) | 5 (15.2) | 0.641 |

| **Variable** | **DBD (n=10,638)** | **DCD (n=33)** | **P value** |
| --- | --- | --- | --- |
| Dialysis, n (%) | 665 (6.3) | 3 (9.1) | 0.740 |
| Stroke, n (%) | 346 (3.3) | 1 (3.0) | 1.000 |
| Permanent pacemaker, n (%) | 92 (0.9) | 0 (0.0) | 1.000 |
| Hospital length of stay (days) | 19.0 [13.0–35.0] | 31.5 [15.5–57.5] | **0.023** |

*Values are median [interquartile range] or n (%). Percentages are calculated from non-missing observations. DBD, donation after brain death; DCD, donation after circulatory death; CMV, cytomegalovirus; HLA, human leukocyte antigen; LV, left ventricular; PLM, predicted left ventricular mass; CNS, central nervous system.*

### Size matching and transplant characteristics

Both donor and recipient left ventricular mass were lower in the DCD group (donor 21.34 vs 45.17 g, P=0.020; recipient 19.07 vs 34.50 g, P=0.004), consistent with the younger age of DCD donors and recipients. Despite these differences, the predicted left ventricular mass ratio was similar (1.36 vs 1.35; P=0.765), and the proportion of transplants achieving a PLM ratio greater than 0.90 did not differ (90.9% vs 92.8%; P=0.941). Weight and height mismatch, HLA mismatch, ischemic time, listing status, and waitlist duration were comparable between groups; donor distance to the transplant center was modestly greater for DCD transplants (465 vs 297 nautical miles; P=0.056).

### Post-transplant outcomes

Early survival was similar between groups at 30 days (100.0% vs 96.1%), 90 days (97.0% vs 93.9%), and 1 year (87.9% vs 90.2%; all P>0.4). Kaplan–Meier survival through 5 years did not differ significantly by donor type (HR 1.17; 95% CI 0.49–2.81; log-rank P=0.73; Figure 2). Rates of treated acute rejection (15.2% vs 18.4%), dialysis (9.1% vs 6.3%), stroke (3.0% vs 3.3%), and permanent pacemaker implantation (0% vs 0.9%) were comparable. DCD recipients had a longer hospital length of stay (median 31.5 vs 19 days; P=0.023).

**Figure 2.**
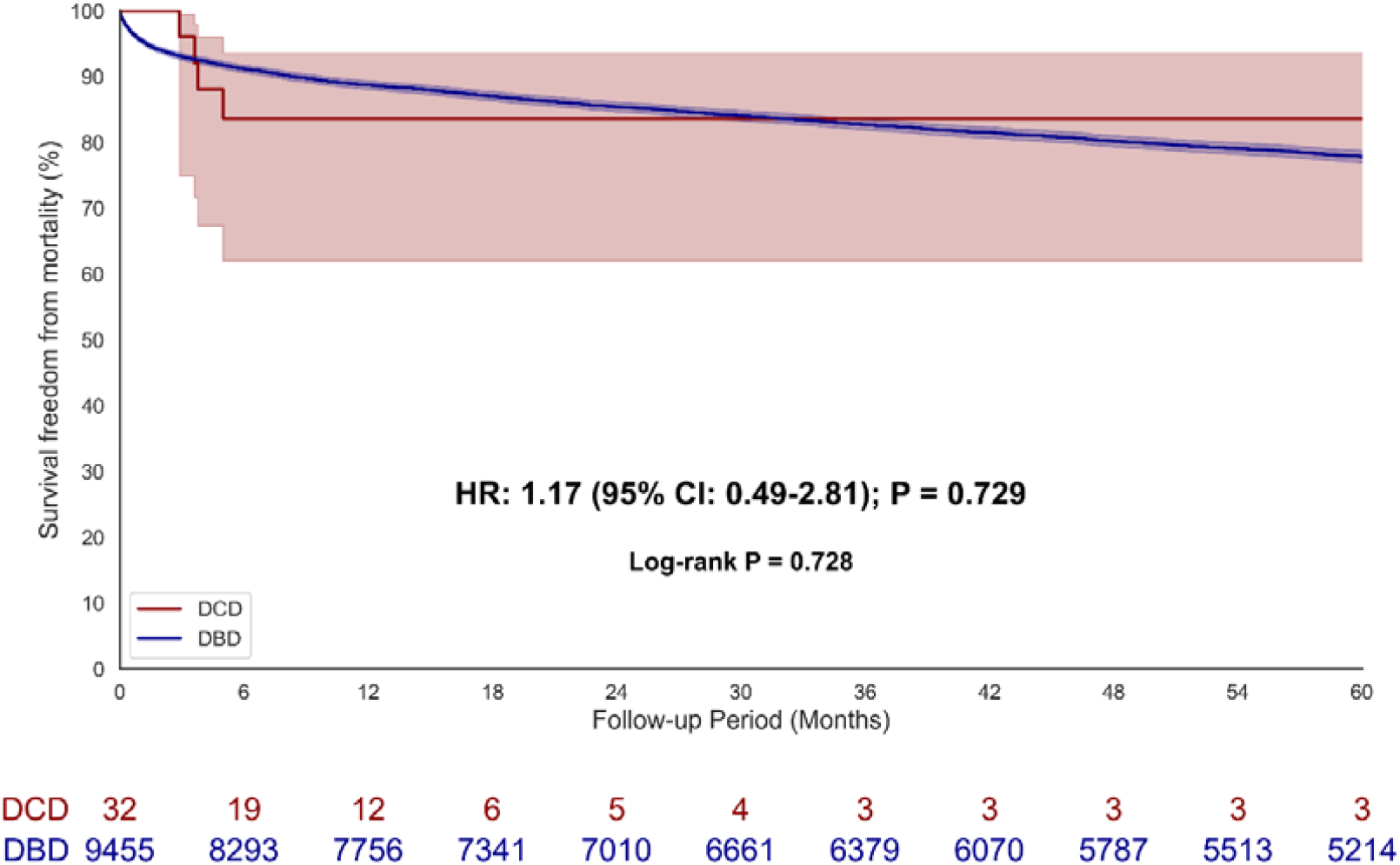
Kaplan–Meier estimates of survival through 5 years after pediatric heart transplantation by donor type. Shaded areas represent 95% confidence intervals. The number at risk is shown below the x-axis. Hazard ratio 1.17 (95% CI 0.49–2.81); log-rank P=0.73.

### Geographic variation

Pediatric DCD transplantation was geographically concentrated: more than 90% of DCD transplants were performed in four UNOS regions—Region 11 (40%), Region 4 (27%), Region 5 (15%), and Region 8 (12%)—while several regions performed none during the study period (Figure 3).

**Figure 3.**
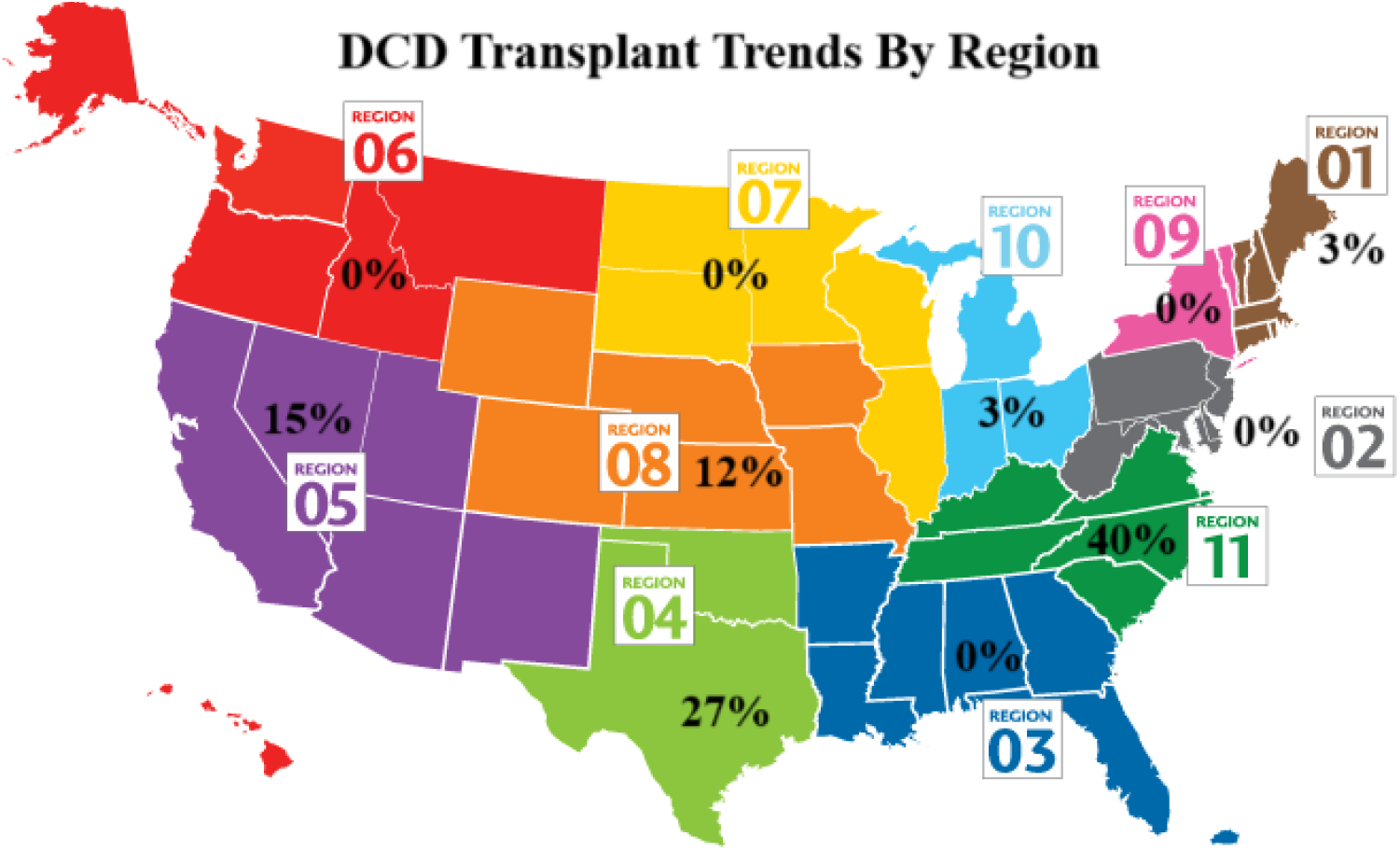
Geographic distribution of pediatric DCD heart transplantation across the 11 UNOS regions, expressed as the percentage of all pediatric DCD transplants performed within each region during the study period.

## Discussion

In this national analysis of more than 10,000 pediatric heart transplant recipients, DCD accounted for approximately 0.3% of transplants but rose sharply in 2023–2024. DCD allografts were used preferentially in infants and children with congenital heart disease, came from younger donors who more often died of anoxia, and—despite lower absolute ventricular mass—achieved size matching comparable to DBD. Early and mid-term survival did not differ significantly between groups, although DCD recipients experienced longer hospitalizations, and DCD activity was concentrated in a small number of geographic regions.

These findings extend adult evidence into the pediatric setting. In adults, the randomized OCS-DCD trial and multiple national analyses have shown that DCD hearts yield survival comparable to DBD while shortening waiting times [5–7], and international series have demonstrated the feasibility of both machine perfusion and in-situ regional perfusion strategies [16]. Our observation of comparable early survival is consistent with these data and contrasts with the earlier pediatric registry experience, in which DCD was associated with lower survival [8]—a difference plausibly attributable to accumulating experience, improved recovery and preservation technology, and more selective application.

Our results are most directly complementary to recent United States registry analyses. A UNOS analysis of pediatric DCD procurement and transplantation before 2024 similarly found DCD to be uncommon but increasing [12], and a contemporary waitlist analysis reported that candidates willing to accept DCD hearts achieved shorter waiting times and lower waitlist mortality without a survival penalty [13]. The present analysis adds a nationally comprehensive, post-transplant comparison that extends through March 2025 and therefore captures the pronounced 2023–2024 surge in utilization, providing a larger and more contemporary DCD cohort than prior pediatric reports. At the same time, reports of worse graft survival among adults with congenital heart disease receiving DCD hearts [14] underscore that outcomes may be population-specific and that ongoing surveillance is warranted as indications broaden.

The predominance of infants and patients with congenital heart disease among DCD recipients is notable. These are precisely the groups with the highest waitlist mortality and the greatest scarcity of size-matched donors [1], suggesting that DCD is being applied where the unmet need is greatest. The preservation of predicted left ventricular mass matching despite lower absolute donor and recipient ventricular mass indicates that appropriate size matching can be maintained in this population. The longer hospital stay observed among DCD recipients may reflect the greater proportion of infants and complex congenital heart disease, the potential for early graft dysfunction after circulatory-death recovery, or center-level practice patterns; the registry does not capture the granular perioperative data needed to distinguish these possibilities.

The marked geographic concentration of pediatric DCD—confined largely to four regions, with many performing none—likely reflects the localization of procurement expertise and perfusion technology, including the emergence of ex-vivo and regional perfusion programs [9–11,16]. This heterogeneity has implications for equity of access: broader dissemination of recovery techniques and standardization of practice could allow more centers and candidates to benefit, and single-center pediatric experiences with ex-vivo systems suggest such expansion is feasible [17].

## Limitations

This study has important limitations. First, only 33 DCD transplants were available, so the analysis is descriptive and hypothesis-generating; comparable outcomes should be interpreted as an absence of a detectable difference rather than proof of equivalence, and the survival comparison is underpowered. Second, DCD transplants were concentrated in 2023–2024 whereas DBD transplants spanned three decades, introducing substantial era imbalance; residual confounding by transplant era cannot be excluded, and the limited number of DCD events precluded robust multivariable adjustment. Third, follow-up for the DCD cohort is short: few DCD recipients contributed to later time points, so 1-year and especially 5-year survival estimates rest on small numbers at risk and should be interpreted cautiously. Fourth, the OPTN registry does not capture procurement strategy (direct procurement with machine perfusion versus normothermic regional perfusion), functional warm ischemic time, or center volume, all of which may influence outcomes. Finally, the analysis is subject to the selection and coding limitations inherent to registry data, including center-level decisions to pursue DCD.

## Conclusions

In this national registry analysis, pediatric DCD heart transplantation was uncommon but expanding rapidly, geographically concentrated, and applied preferentially to infants and children with congenital heart disease. Early and mid-term post-transplant outcomes were not significantly different from DBD, and appropriate size matching was maintained. These findings support the cautious, monitored expansion of DCD as a strategy to enlarge the pediatric donor pool, particularly in currently underutilized regions, while larger cohorts with longer follow-up are needed to confirm long-term safety and to define optimal recipient selection.

## Author Contributions

B.K.M. and R.G. (co-first authors) contributed equally and were responsible for conceptualization, methodology, formal analysis, data curation, and writing of the original draft. O.A.K., G.M., P.D., Y.P.S and N.K.A. contributed to data curation, investigation, and writing—review and editing. J.K. contributed to methodology and critical revision. S.S. was responsible for conceptualization, supervision, and writing—review and editing. B.K.M. served as corresponding author. All authors have seen and approved the final manuscript and agree to be accountable for all aspects of the work.

## Declarations

### Funding

The authors received no specific funding for this work.

### Competing interests

The authors declare no competing interests.

### Ethics approval

This study used de-identified data from the OPTN/UNOS registry and was determined to be exempt from institutional review board review; the study conformed to the principles of the Declaration of Helsinki.

### Consent

Not applicable; the analysis used de-identified registry data.

### Data availability

The data underlying this article were obtained from the OPTN/UNOS Standard Transplant Analysis and Research (STAR) files. These data are available to qualified investigators through the OPTN and cannot be redistributed by the authors owing to the terms of the data use agreement.

### Prior presentation

Presented at the Eastern Cardiothoracic Surgical Society (ECTSS) Annual Meeting, 2025 (Abstract C02).

## Acknowledgements

This work was supported in part by Health Resources and Services Administration (HRSA) contract [contract number]. The data reported here have been supplied by the United Network for Organ Sharing (UNOS) as the contractor for the OPTN. The interpretation and reporting of these data are the responsibility of the authors and in no way should be seen as an official policy of, or interpretation by, the OPTN or the U.S. Government.

## Notes

**Conflicts of interest:** The authors declare no conflicts of interest.

### Competing Interest Statement

The authors have declared no competing interest.

### Author Declarations

The Institutional Review Board of the University of Miami waived ethical approval for this work.

